# Image-based modelling of inhaler deposition during respiratory exacerbation

**DOI:** 10.1101/2020.06.08.20118513

**Authors:** Josh Williams, Jari Kolehmainen, Steve Cunningham, Ali Ozel, Uwe Wolfram

**Affiliations:** School of Engineering and Physical Sciences, Heriot-Watt University, Edinburgh, UK; Department of Chemical and Biological Engineering, Princeton University, Princeton, New Jersey, USA; Centre for Inflammation Research, University of Edinburgh, Edinburgh, UK

**Keywords:** Aerosol deposition, Respiratory exacerbation, Patient-specific modelling, Drug dosimetry, Metered-dose inhaler, Computational fluid dynamics

## Abstract

For many of the one billion sufferers of respiratory diseases worldwide, managing their disease with inhalers improves their ability to breathe. Poor disease management and rising pollution can trigger exacerbations which require urgent relief. Higher drug deposition in the throat instead of the lungs limits the impact on patient symptoms. To optimise delivery to the lung, patient-specific computational studies of aerosol inhalation can be used. However in many studies, inhalation modelling does not represent an exacerbation, where the patient’s breath is much faster and shorter. Here we compare differences in deposition of inhaler particles (10, 4 µm) in the airways of a healthy male, female lung cancer and child cystic fibrosis patient. We aimed to evaluate deposition differences during an exacerbation compared to healthy breathing with image-based healthy and diseased patient models. We found that the ratio of drug in the lower to upper lobes was 35% larger during healthy breathing than an exacerbation. For smaller particles the upper airway deposition was similar in all patients, but local deposition hotspots differed in size, location and intensity. Our results identify that image-based airways must be used in respiratory modelling. Various inhalation profiles should be tested for optimal prediction of inhaler deposition.

**Highlights:**

- Regional and local drug deposition was modelled in three patients during normal, sinusoidal inhalation and an exacerbation.
- Local drug deposition changes with airway shape and inhalation profile, even when regional deposition is similar.
- Image-based models were combined with highly-resolved particle tracking including particle contact and cohesion.
- Fluid model validated by comparing gas velocity field with in vitro experiments.

## 1. Introduction

More than one billion people worldwide suffer from asthma, cystic fibrosis, and other chronic respiratory diseases (The Global Asthma Network, 2018), many experiencing distress and anxiety due to restrictions to activities and limited productivity (Dockrell et al., 2007). These limitations are most prominent among the young and elderly (The Global Asthma Network, 2018). One of the largest contributors to the diseased population is asthma, which incurs an annual cost per patient of €1,700 and $3,100 in Europe and the USA, respectively (Nunes et al., 2017) from direct cost of treatment and indirect costs such as work absence or decreased productivity (Katsaounou et al., 2018; Gruffydd-Jones et al., 2019). Similar impacts are induced from cystic fibrosis (Chevreul et al., 2015). Consistent treatment in alignment with disease management plans are recommended to minimise symptoms (Ring et al., 2015), but adherence is an issue in young patients (McQuaid et al., 2003) and many adults are purposely inconsistent to limit exposure to side-effects such as osteoporosis and cataracts (Dockrell et al., 2007). Even in adherent patients, efficiency of the metered-dose inhaler varies greatly across patients (Clark, 1995) as many (particularly children) experience difficulties in device technique (Usmani, 2019) due to the rapid spray of the drug. The issue of technique (patient breathing pattern and coordination with device actuation) and differences in lung structure are the main influences in drug delivery (Darquenne et al., 2016).

Optimisation of the medication deposition could be achieved through *in silico* analyses, by providing the clinician information on the local deposition and therapeutic outcome. One available deposition tool is the Multiple-Path Particle Dosimetry (MPPD) model (Anjilvel and Asgharian, 1995; Asgharian et al., 2001). However this calculates deposition of ambient particles (Borghardt et al., 2015), which does not mirror the physics of spray aerosol inhalation (Longest et al., 2008). This issue has been recognised and a commercial counterpart to predict deposition of pharmaceutical aerosols has been developed (Olsson and Bäckman, 2018). However, as the equations used are based on probabilistic 1D equations, similar to that of MPPD, complex fluid phenomena generated in the upper airways and particle interactions cannot be included. Comparisons between 1D models and computational particle-fluid dynamics (CPFD) deposition led Zhang et al. (2009) to observe significant differences in local deposition due to local flow features not captured in 1D models. Flow and particle phenomena can be readily accounted for in CPFD by solving equations governing the transport of air and particles (e.g. see Sundaresan et al. (2018)). Flow can be solved in airways extracted from medical images and particle properties can represent inhaler particles to produce patient-specific deposition analyses. But with the complexity of the flow regimes in the system creating demanding simulations and the massive number of respiratory patients, the benefit-to-cost of image-based modelling is an issue. Work exists interpreting pharmaceutical deposition differences in adult patients (Feng et al., 2018; van Holsbeke et al., 2018; Poorbahrami and Oakes, 2019; Poorbahrami et al., 2019), but child deposition models are limited to idealised geometries (Das et al., 2018; Longest et al., 2006; Deng et al., 2018). Existing children-focused image-based deposition studies analyse the nasal cavity (Xi et al., 2011, 2012) or central airways (Oakes et al., 2018), which neglects the effect of turbulence generated in the mouth and throat. To move towards enhanced treatments for all ages, a study considering the impact of the upper airways is needed. We begin this by comparison of deposition in the airways (from mouth to central airways) in a group of three patients, containing a child and two adults. This can allow simulation of flow created in the upper airways and provide understanding of how this affects deposition throughout the airways, across patients.

Results from these simulations are dependent on the inflow conditions, which here is based on the duration and strength of the patient’s inhalation. Colasanti et al. (2004) showed the breathing profile of a patient with extremely obstructed airways, which differed in shape and magnitude from the sinusoidal inhalation waveform that is typically used in respiratory CPFD studies (Inthavong et al., 2010). A sinusoidal inhalation waveform is used by most existing studies (Oakes et al., 2018; Inthavong et al., 2010; Naseri et al., 2017), which mimics a healthy patient’s tidal breathing. The variation in flow patterns produced by these differences would therefore give a deposition analysis which may not represent the flow during bronchoconstriction, when a ‘reliever’ inhaler (bronchodilator) is used. Different breathing profiles have been utilised (Longest et al., 2012; Khajeh-Hosseini-Dalasm and Longest, 2015), but these were used to represent techniques for different devices (dry-powder compared to metered-dose inhalers), not exacerbating and healthy breathing in a metered-dose inhaler. By applying different inflow conditions, one can understand changes in flow structure during an exacerbation or another desired breathing situation. This knowledge would allow manufacturers to tailor inhaled therapeutics to ensure optimal dosage reaches the desired site in the airways, and to provide clinicians and patients the tools with which to maximise inhalation technique during exacerbations.

In addition, existing MDI deposition research simplifies particle interactions, which includes treating particle-wall collision as complete sticking. This is a coarse approximation of dissipative lubrication forces between the particle and wall (Legendre et al., 2005; Holbrook and Longest, 2013). To treat as fully sticking therefore neglects rebound and could over-estimate deposition of high inertia particles. Furthermore, MDI studies often exclude van der Waals forces (Hamaker, 1937) which can cause small particles agglomerate, and become more inertial. If the increase in inertia is large enough, this can change particle trajectory or chance of rebound, which, in turn would cause early deposition. How these forces alter deposition and compete against each other should be tested. Applying this to simulations made specific to a patient’s airway structure under extreme and optimal breathing will further understanding of drug transport across different health states.

Therefore, we aimed to (i) evaluate the effect of patient-specific airway shape on drug deposition. To satisfy this we also aimed to (ii) identify the necessary physical effects to produce accurate models of the system (including the mass of drug simulated, van der Waals and particle-wall lubrication forces), and (iii) evaluate change in deposition during an exacerbation compared to healthy breathing.

## 2. Methods

To answer the research aims above, we evaluated changes in deposition produced by the following variations. Parameters varied included the mass of drug simulated, particle cohesiveness, lubrication forces in particle wall-collisions, and deletion or saving of deposited particles from the system to mimic absorbing particles with mucus layer. The use of a healthy and exacerbation breathing profile (Colasanti et al., 2004) allowed for analysis of the reliever inhaler during an exacerbation. The effects of patient variation were then analysed using this optimised modelling setup, through comparison under realistic inflow conditions in three patients.

### 2.1. Medical image processing

Three patients were studied retrospectively using computed tomography (CT) (detailed in Table 1). Use of these retrospective images was approved by Heriot-Watt University (ID: 2020-0500-1452). The surface file of the healthy patient’s segmented airways was provided by the study of Banko et al. (2015). The patients had sufficient variation in age, gender and health status to gather an indication of the benefit of patient-specificity.

**Table 1:** Information of patients studied in this work. Although a small group, variations in gender, age and disease have been included.

| Gender | Age | Illness | Voxel size [mm<sup>3</sup>] | Source |
| --- | --- | --- | --- | --- |
| Male | 45–50 | Healthy | $0.977 \times 0.977 \times 2.5$ | Zhang et al. (2012),<br>Banko et al. (2015) |
| Female | Unknown (adult) | Lung cancer | $0.977 \times 0.977 \times 3.0$ | Yang et al. (2017)<br>Yang et al. (2018)<br>Clark et al. (2013) |
| Male | 10–15 | Cystic fibrosis | $0.5254 \times 0.5254 \times 1.0$ | |

Images were processed using 3D Slicer 4.10 (Fedorov et al., 2012). Images were preprocessed by applying isotropic spacing to account for the anisotropic resolution of CT scans (Table 1). For the cancer patient, voxel size was then reduced using a linear interpolation, creating a voxel size of ≈ 0.4 mm to allow extraction of smaller airways. This was also performed on the cystic fibrosis patient. To preserve edges of the airway while blurring lung tissue, an anisotropic diffusion filter was then applied (Duan et al., 2019). Parameters used were conductance 3, 5 iterations and step size of 0.06025. These were based on visual comparison of our segmentations produced from varying values of conductance used by Behnaz et al. (2010) and Sen et al. (2011).

Images were then segmented using a threshold-based region-growing approach to segment areas classified as air without leakage to the background air (Nardelli et al., 2015; Mayer et al., 2004; De Nunzio et al., 2011; Aykac et al., 2003). This semi-automatic process grows the segment from ‘seeds’ which are user declared points within each region, set separately for the airway and surrounding lung tissue. Once each region was grown, labels denoting leaked regions were found by overlaying the segmentation on the scan. Leaked regions were then labelled as airway and the region was grown again until the scan was observed to be acceptable quality. This process was applied to the external airways, right and left lungs separately due to variations in the airway image density in each region (Nardelli et al., 2015). This allowed segmentation to a depth ranging from the fourth to sixth bifurcation level (G4 – G6, Figure 1).

**Figure 1:**
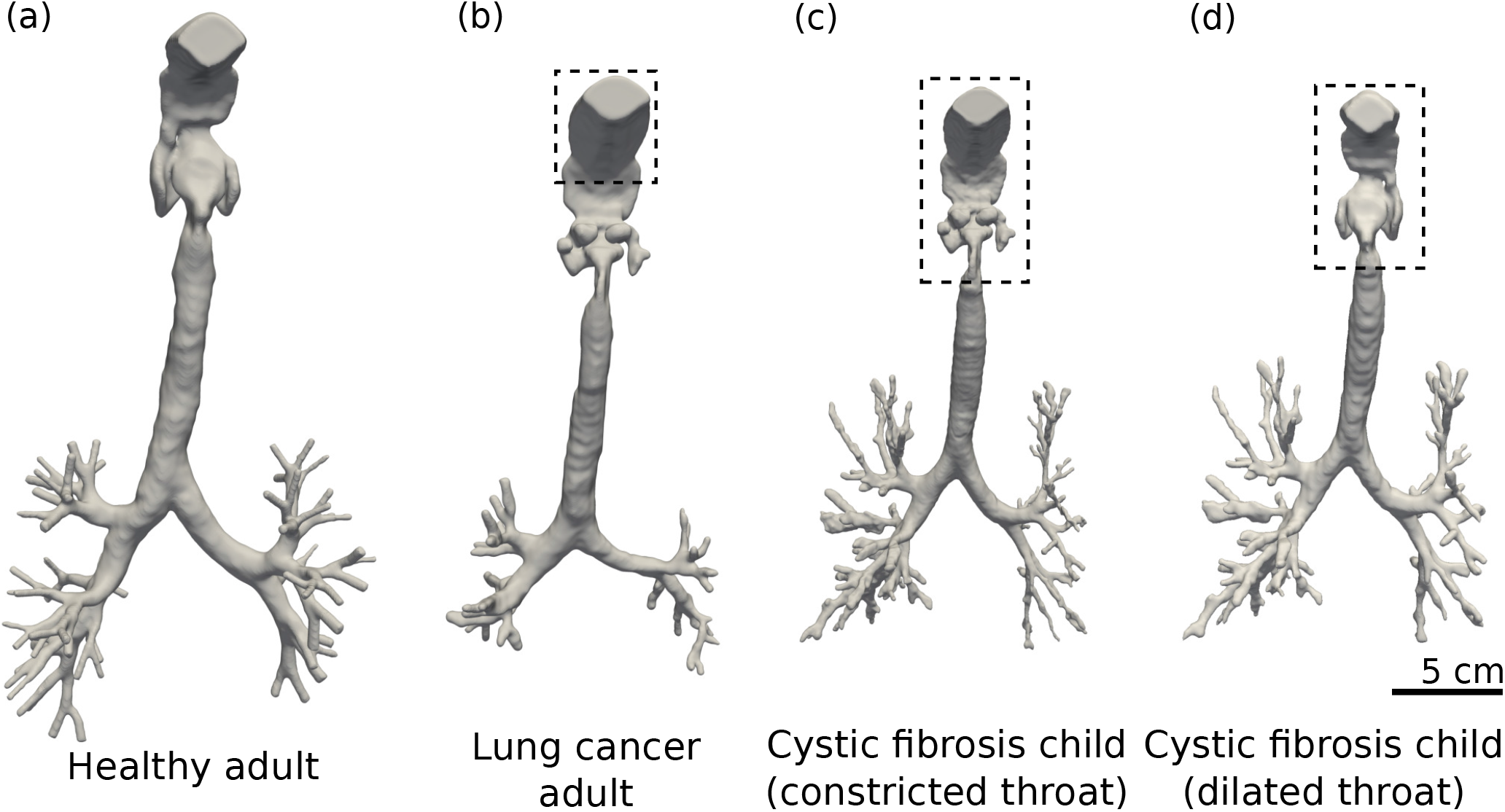
Segmented airway trees of patients included in the study. The airways are presented from left to right as (a) the healthy, male adult (Banko et al., 2015), (b) the adult, female lung cancer patient (Yang et al., 2017, 2018; Clark et al., 2013), and (c,d) the male child cystic fibrosis patient. The child patient has (c) constricted throat added from the cancer patient (b), and (d) has a dilated throat taken from the healthy patient (a). Areas within the dashed box have been artificially added due to available images not including this region.

Some of the upper airways were not included in the CT data, we have found this to be common in most clinical CT scans. To account for this we merged the oral cavity of the healthy patient to the throat of the lung cancer patient. This region was then extracted, scaled and joined to the trachea of the cystic fibrosis patient to complete the missing regions. Scaling was performed such that the intersection of the new and existing regions matched in diameter (resulting in a scaling of 0.8 for the added region). This was later repeated using the throat of healthy patient, after being advised such a narrow, obstructed throat is not characteristic of cystic fibrosis, and likely unique to the cancer patient. The regions which were artificially added are shown graphically in Figure 1 within the dashed box. These excluded regions are of large importance in inhaler simulations as a large portion of the dosage is lost within this part of the airway and the turbulence created here is cascaded through the trachea and main bronchus (Banko et al., 2015).

### 2.2. Mathematical modelling

Here we present the mathematical relations used to determine the physical factors included in our model of the system. We provide the equations governing the fluid and particle solvers in Appendix A. Briefly, particle transport was solved by the discrete element method (DEM) using the particle simulator LIGGGHTS (Kloss and Goniva, 2011). This tracks each individual particle’s trajectory by integrating Newton’s equations of motion in time (Verlet, 1967). Particle collisions were modelled as a linear spring-dashpot system (Cundall and Strack, 1979). Fluid transport through the airways was solved by the volume-filtered mass and momentum conservation equations (Anderson and Jackson, 1967; Capecelatro and Desjardins, 2013) implemented in OpenFOAM v2.2 (Weller et al., 1998). Particle and fluid phases were coupled through a version of the CFDEMcoupling platform (Kloss et al., 2012) modified to benefit from faster two-way coupling by Ozel et al. (2016).

Filtering the fluid transport equations creates unresolved stresses. In our simulations, we consider residual stresses from volume-filtering of fluid velocity fluctuations (***R***_*u*_) below the cell size, Δ. ***R***_*u*_ is dependent upon the eddy viscosity (*µ*_*t*_), a term representing turbulence dissipation into the smaller, unresolved scales. Eddy viscosity is modelled using a dynamic Smagorinsky model (Germano et al., 1991; Lilly, 1992). This uses a second filter with width 2Δ to sample the smallest resolved scales, and calculate the local Smagorinsky constant *C*_*S*_. Using the classic Smagorinsky (1963) model requires a global *C*_*S*_ which makes the model unable to represent a range of flow (Germano et al., 1991), such as the laminar-turbulent-laminar transitions in the airways.

We compute drag forces acting on particles using Beetstra et al. (2007)’s model for monodisperse particles. This drag model is based on the particle’s Reynolds number, *Re*_*p*_, given as

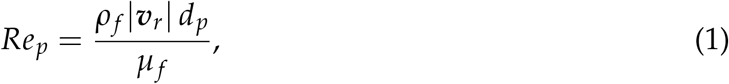

where |***υ***_*r*_| is the magnitude of the relative velocity vector between gas and particle velocities, *d*_*p*_ is the particle diameter, *µ*_*f*_ is the molecular viscosity of the gas phase.

As well as the particle Reynolds number, we also characterise our system through the particle’s Stokes number, *St*. This is the ratio of particle timescale (*τ*_*p*_) to fluid timescale (*τ*_*f*_), given by

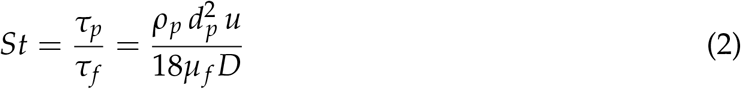

where *D* is the inlet diameter, and we take *u* as the peak cross-sectional velocity at the inlet Kleinstreuer and Zhang (2003). This gives *St* >> 1 when particles have high inertia and are not perturbed by fluid fluctuations. Conversely, when *St* << 1 the particles follow the fluid streamlines closely and will have lower deposition in the upper airways. In gas-solid flows, when *St* ≈ 1 particles will concentrate in clusters of low fluid vorticity (Squires and Eaton, 1991).

Again, due to the particle’s small size, particle-particle cohesion from van der Waals forces may influence deposition. Particle attractive energy due to van der Waals force is determined by the material’s Hamaker constant, *A* (Hamaker, 1937). For a linear sliding-dashpot particle-particle collision model, it is not affordable to use a realistic stiffness due to limitation of very small time-step. Therefore, to permit a larger timestep, the elastic properties of the particles were softened. Therefore the real stiffness (*k*_*R*_) is reduced to a soft stiffness (*k*_*S*_), and the Hamaker constant is amended by the relationship *A*^*S*^ = *A*^*R*^(*k*_*S*_/*k*_*R*_)^1/2^ using the model of Gu et al. (2016*a*). This reduction of real particle stiffness (*k*_*R*_) to a softer stiffness (*k*_*S*_) has negligible effect on fluid hydrodynamics and particle cohesion (Gu et al., 2016*a*; Ozel et al., 2017). To determine *A*, which is not given in literature for the metered-dose inhaler propellant HFA-134A, we evaluate deposition at three Bond numbers, *Bo. Bo* is provided by Ozel et al. (2017) as

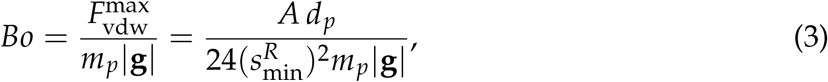

where 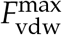 is the maximum van der Waals force magnitude occuring when 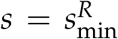 (the minimum separation distance for a particle at its real stiffness). This was varied three orders of magnitude, *Bo* = 10, 100, 1000 which was sufficient variation to interpret cohesive differences. This magnitude of variation was chosen as it showed changes in deposition without running a large amount of simulations at finer *Bo* intervals. This also gave *A* at *Bo* = 1000 of the same order of magnitude (*A* = 10^−20^ J) to drug particles in the inhaler propellant HFA-227 (Engstrom et al., 2009).

Further complexities arise when considering particle-wall interactions. This is widely treated as a fully plastic collision due to the presence of a respiratory mucus layer (Miyawaki et al., 2012; Zhang et al., 2018; Chen et al., 2012; Naseri et al., 2017). This may occur due to lubrication interactions, which have been experimentally shown to damp collision forces (Legendre et al., 2005) due to the formation of a thin interfacial film during contact. This relation has been shown to follow the expression

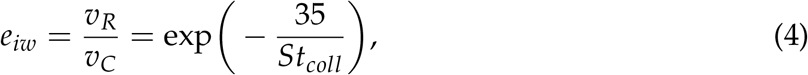

for solid particles (Legendre et al., 2005). Here, subscripts (.)_*R*_ and (.)_*C*_ are the rebound and pre-collision velocities, respectively, *υ*_*T*_ is the velocity of the particle before collision and the collision Stoke’s number (Legendre et al., 2006) is given as

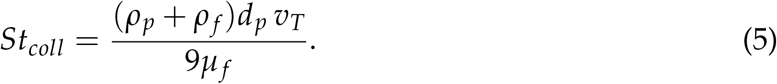

The effective restitution coefficient, *e*, follows a sigmoid trend when plotted against *St*_*coll*_, with *St*_*coll*_ < 10 creating a plastic collision similar to that approximated in modelling of particle-mucus layer interactions. We implemented this relationship to model particle-wall contact force, then when later deleting or freezing deposited particles, use it to determine the cutoff. We use this to model particle-mucus layer collision instead of the typical ‘sticking’ condition (Miyawaki et al., 2012; Zhang et al., 2018; Chen et al., 2012; Naseri et al., 2017). This allows potential for particle rebound after impacting the wall. Therefore, as the particle-wall interaction is better represented, deposition is not over-estimated.

### 2.3. Simulation configuration

#### 2.3.1. Fluid-phase Simulations

We first validated the fluid phase by comparison to a published experimental study (Banko et al., 2015). The study observed water flow in a 3D printed hollow cast of an adult male patient’s airways (Figure 1a) at a Reynolds number representative of heavy breathing (*Re*_inlet_ = *ρ* _*f*_ *UD*_inlet_/*µ*_*f*_ = 3600 corresponding to *Q* = 1 L/s). First we simulated water flow through the airways to validate our fluid solver. We then simulated airflow at the same *Re*. Single-phase (water) transport through lung airways was simulated using OpenFOAM solver pimpleFoam, then when simulating air we used the particle-carrier phase, or namely CPFD, solver with no particles. We investigated the effect of mesh resolution on the results by simulating two uniform, hexahedral meshes (5 × 10^5^ and 10^6^ cells). We used the mesh with 10^6^ cells for all simulations after the mesh sensitivity study. As interaction between the gas and particle phases (namely, drag) is dependent on the size of Δ relative to *d*_*p*_ (Agrawal et al., 2001), we made the size of cell in our 10^6^ mesh Δ = 500 µm = 50*d*_*p*_ to minimise drag overestimation. This is of the same order of magnitude as coarse-fluid grid simulations performed by Radl and Sundaresan (2014). We kept grid size consistent with our first particle size tested (at *d*_*p*_ = 10 µm, Δ = 500 µm) throughout the study to minimise excessive computation time when later reducing particle size.

We applied an inlet condition which added random components to the inlet flow velocity (Sagaut, 2006) to account for upstream fluctuations in the experimental apparatus. This was used with an inflow velocity of *u*_water_ = 0.167 m/s and *u*_air_ = 2.677 m/s, with a no-slip condition at the wall. Outlets had a uniform, fixed pressure applied to each bronchi. Although not truly representative of outflow conditions within the lung, a uniform outlet condition has been shown to be acceptable when the bronchi are extended a few diameters in the axial direction to eliminate secondary flow effects (Zhang et al., 2012). This has been shown to resemble more advanced outlet conditions which consider compliance of the lung (Ma and Lutchen, 2006).

#### 2.3.2. Aerosol transport simulations

In multiphase simulations we coupled the single-phase approach above to the DEM solver to track monodisperse particles of *d*_*p*_ = 10 and 4 µm. Particles are classed as deposited and deleted from the system when impacting the wall with low inertia (*St*_*coll*_ < 20). As CT scan resolution only permitted segmentation to approximately the sixth bifurcation level (Figure 1), particles reaching the end of the bronchial path were classified as reaching the distal airways and deleted. As we only model a limited amount of bifurcation levels (between four to six), this is a coarse approximation of downstream behaviour and dosage reaching the small airways.

The dosage was released over a period of *t* = 0.1 s (Ju et al., 2010). The particle motion was discretised in time based on calculation of the collision time of the particles, 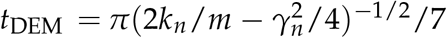 (Gu et al., 2016*a*). The van der Waals stiffness scaling (Gu et al., 2016*a*) was set to *A*^*S*^ = *A*^*R*^(*k*_*S*_/*k*_*R*_)^1/2^ = *A*^*R*^/31.6. Particle lubrication force was modelled using mucus layer viscosity ranging from *µ*_mucus_ = 0.026 − 0.05 Pa · s dependent on disease (Rubin, 2007) and a density of *ρ*_mucus_ = 1000 kg/m^3^ as it is largely made up of water in the upper airways (Olsson et al., 2011). Parameters used are summarised in Table 2. Simulations were performed on high-performance computers ROCKS of Heriot-Watt University and EDDIE of the University of Edinburgh high-performance computers using 28 – 64 CPU cores. Simulations with 10 µm particles took 2-6 weeks. Simulations with 4 µm particles took 4-16 weeks.

**Table 2:**
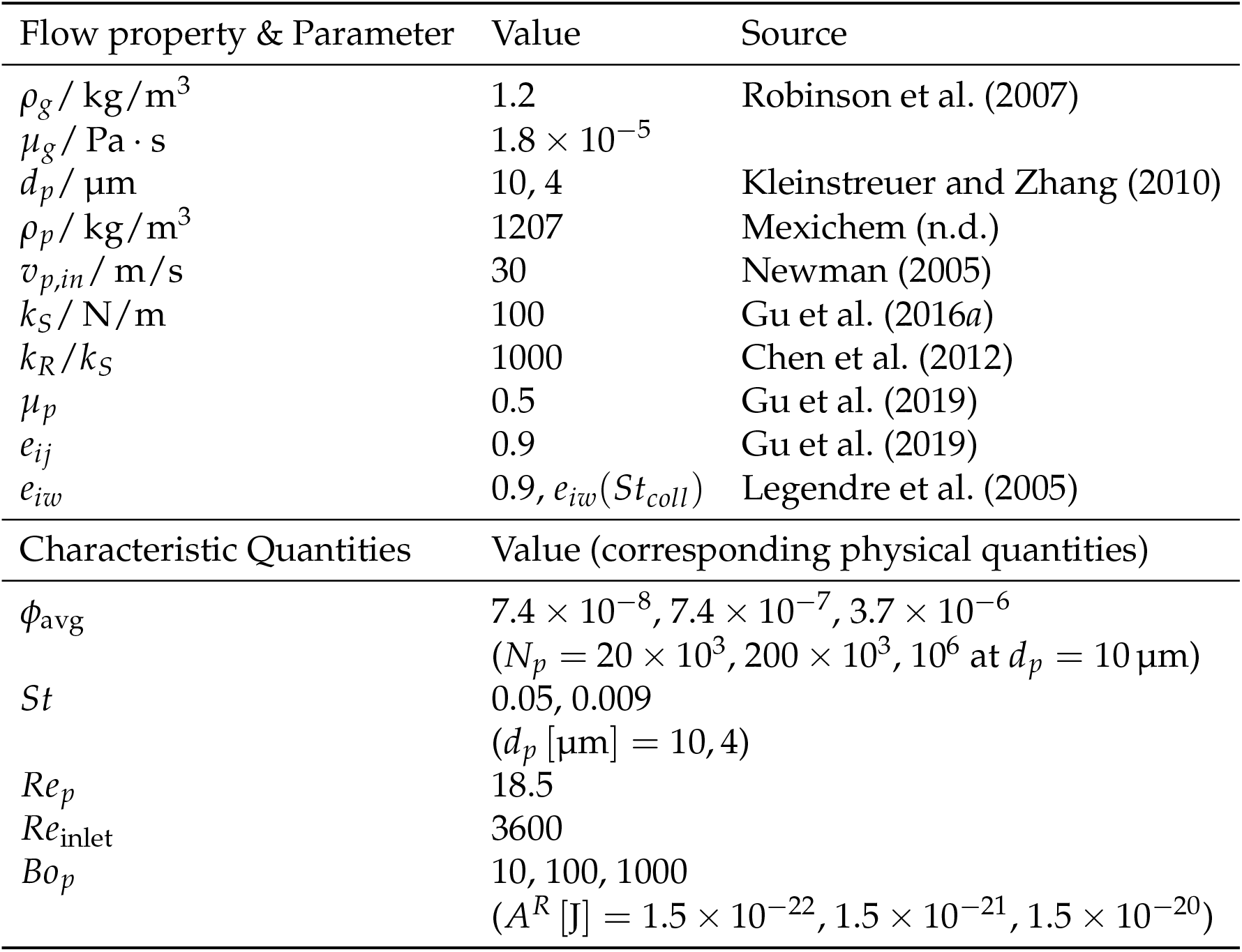
Flow properties with model parameters used in simulations, as well as dimensionless quantities varied. Three separate volume fractions were used to observe sensitivity to the number of dosage simulated. Three Bond numbers were simulated to determine the effect of van der Waals forces. Finally, particle-wall lubrication forces were included by comparing a collision with fixed restitution coefficient *e* and one dependent on *St*_*coll*_.

| Flow property & Parameter | Value | Source |
| --- | --- | --- |
| $\rho_g / \text{kg/m}^3$ | 1.2 | Robinson et al. (2007) |
| $\mu_g / \text{Pa} \cdot \text{s}$ | $1.8 \times 10^{-5}$ | |
| $d_p / \mu\text{m}$ | 10, 4 | Kleinstreuer and Zhang (2010) |
| $\rho_p / \text{kg/m}^3$ | 1207 | Mexichem (n.d.) |
| $v_{p,in} / \text{m/s}$ | 30 | Newman (2005) |
| $k_S / \text{N/m}$ | 100 | Gu et al. (2016a) |
| $k_R / k_S$ | 1000 | Chen et al. (2012) |
| $\mu_p$ | 0.5 | Gu et al. (2019) |
| $e_{ij}$ | 0.9 | Gu et al. (2019) |
| $e_{iw}$ | $0.9, e_{iw}(St_{coll})$ | Legendre et al. (2005) |
| Characteristic Quantities | Value (corresponding physical quantities) |  |
| $\phi_{avg}$ | $7.4 \times 10^{-8}, 7.4 \times 10^{-7}, 3.7 \times 10^{-6}$<br>($N_p = 20 \times 10^3, 200 \times 10^3, 10^6$ at $d_p = 10 \mu\text{m}$) | |
| $St$ | 0.05, 0.009<br>($d_p [\mu\text{m}] = 10, 4$) | |
| $Re_p$ | 18.5 | |
| $Re_{inlet}$ | 3600 | |
| $Bo_p$ | 10, 100, 1000<br>($A^R [\text{J}] = 1.5 \times 10^{-22}, 1.5 \times 10^{-21}, 1.5 \times 10^{-20}$) | |

Once the optimal parameters were determined, a time-varying inlet condition was implemented to represent a real breathing cycle (Figure 2). For this we used data provided by Colasanti et al. (2004) whom analysed the breathing profile of patients suffering from COPD and cystic fibrosis with extreme airway obstructions. Colasanti et al. (2004) also presented a healthy patient’s inhalation profile. The magnitude of the child’s inhalation velocity was lowered by half, based on inhalation velocities used in a similar study (Longest et al., 2006). To compare the effect of inhalation waveform, all patients were simulated using both the healthy and exacerbation profile (Figure 2) using 10 µm and 4 µm particles. To evaluate the effect of the missing throat and mouth of the cystic fibrosis patient, we model this patient using the mouth and throat of the healthy patient and cancer patient.

**Figure 2:**
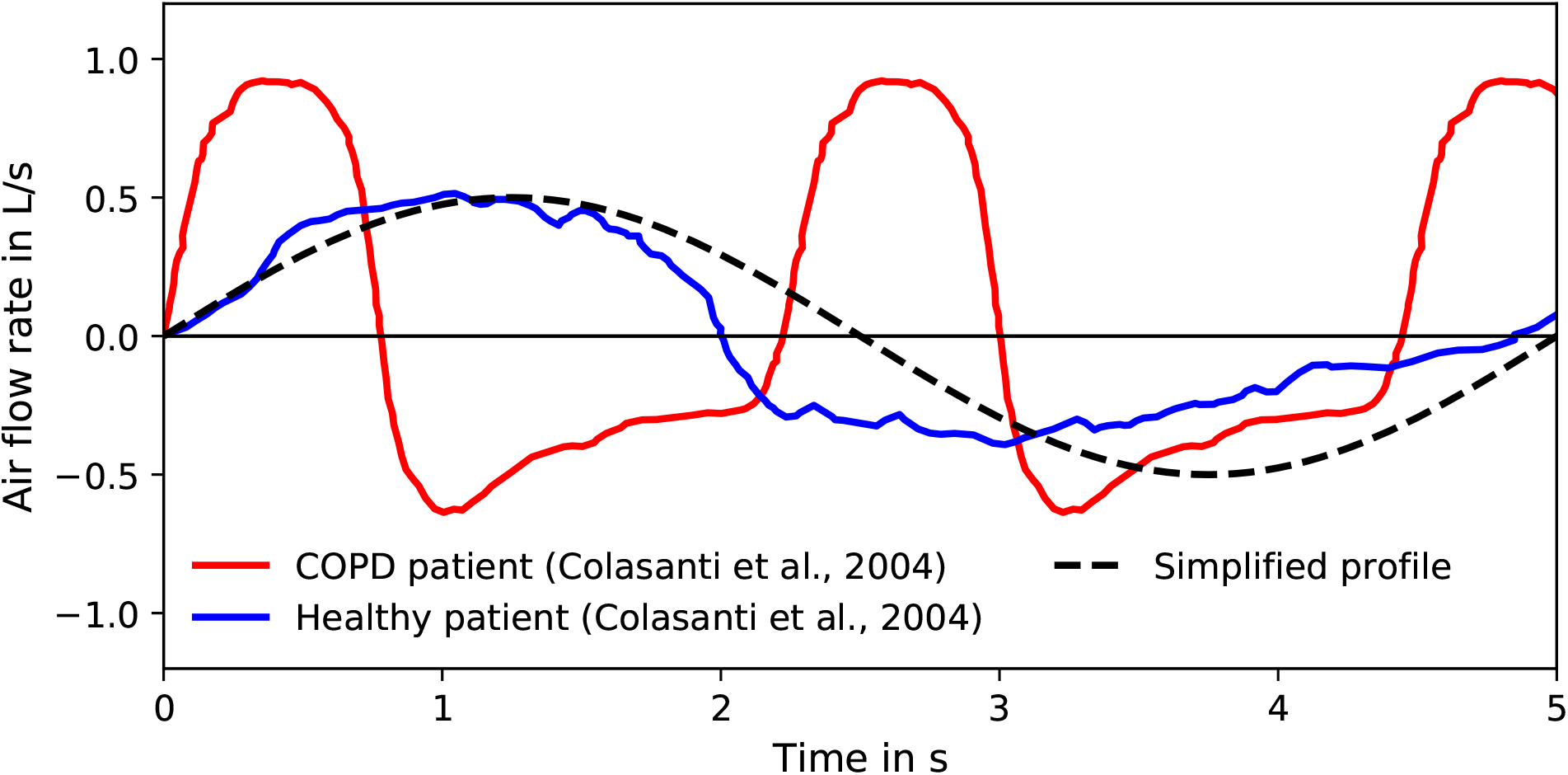
Breathing profiles for healthy and diseased patients used in this study (Colasanti et al., 2004). Positive flow rate represents inhalation and negative represents exhalation. A standard sinusoidal breathing profile is shown for comparison. Differences between healthy and simplified breathing conditions are minor, but the exacerbation profile reaches a higher peak inhalation at a faster rate. Adapted from Williams et al. (2021) with permission of RDD Online LLC.

### 2.4. Deposition evaluation

Particle deposition was analysed by regional groupings of the mouth, throat, trachea, main bronchus and bronchi within each lobe of the lung (as used by Asgharian et al. (2001) and van Holsbeke et al. (2018)). We also provided results grouped into external airways (mouth to end of first branch), and internal airways (airways within the lungs). We use this to evaluate preferential deposition within airways’ different regions.

In contrast to the fully plastic (‘sticking’) particle-wall condition used in literature, our particles may rebound as we resolve collisions over multiple timesteps. We classed particles that had low inertia prior to impacting the wall as deposited. This inertia was based on Equation (4). We compared results where these particles were kept active in the domain to interact with floating particles, or where we simply deleted them. The difference in these was found to be minimal (under 5% in all regions). Therefore we opted to delete them due to superior computational efficiency (reached 0.1 s physical time in 30% faster clock time).

Due to the particle’s ability to slide along the wall in our parameter study simulations, when stuck it would not be completely stationary. To extract particles on the wall for comparison to our sticking condition, deposition was defined when the particle velocity was sufficiently below that of the free-stream (*υ* = 0.01 m/s, 900 times less than gas velocity in the throat). This velocity cutoff was found through observed comparison of velocities of slowly floating and deposited sliding particles. Particles that were below this threshold were classed as deposited deleted during post-processing.

Due to the locally-acting nature of inhalers (Lu et al., 2015), it is important to understand therapeutic distribution and dosage experienced by the patient based on deposition concentration (Solomon et al., 2012). We interpreted this through the dosimetry measure of deposition enhancement factor (DEF) (Balashazy et al., 1999; Longest et al., 2006). We calculate this using the number of particles deposited within a fixed distance (1 mm, area *A*_*conc*_ = *π* (1 mm)^2^) of the central point of each wall face. This distance was based on that used by Dong et al. (2019). Other studies have used much narrower radii (Longest et al., 2006; Xi et al., 2012), but using this wider radius can account for particle translocation during the time between deposition and absorption. This is made relative to the global deposition by

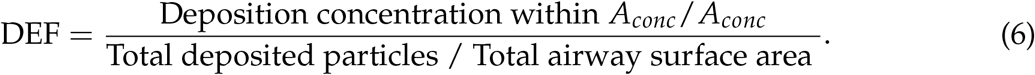

We use this instead of the sum of the deposition efficiencies as the defined areas may overlap, and this therefore prevents particles being counted multiple times towards the global average.

## 3. Results

### 3.1. Validation of Carrier-phase Simulations

As discussed before, dynamics of low-inertia aerosols (as *St* < 1) is significantly altered by carrier-phase flow field. Hence, we first focused on carrier-phase flowfield validation. Turbulence induced in the upper airways is the foundation of the flow structure, creating secondary flows which are responsible for deposition in the trachea (Jin et al., 2007; Kleinstreuer and Zhang, 2010). Therefore, the upper airways are a suitable region for the validation (Figure 3). All results have been normalised by the mean velocity in the trachea (*V*_*T*,water_ = 0.22 m/s and *V*_*T*,air_ = 3.51 m/s). The strong jet of flow formed at the throat matches the experimental study well in magnitude and structure, capturing the recirculation zones as the throat expands well. Banko et al. (2015) gave the bulk (area-averaged) velocity *U* relative to *V*_*T*_ as 1.73 in the glottis, compared to our value of 1.77, producing a 2.3% relative error.

**Figure 3:**
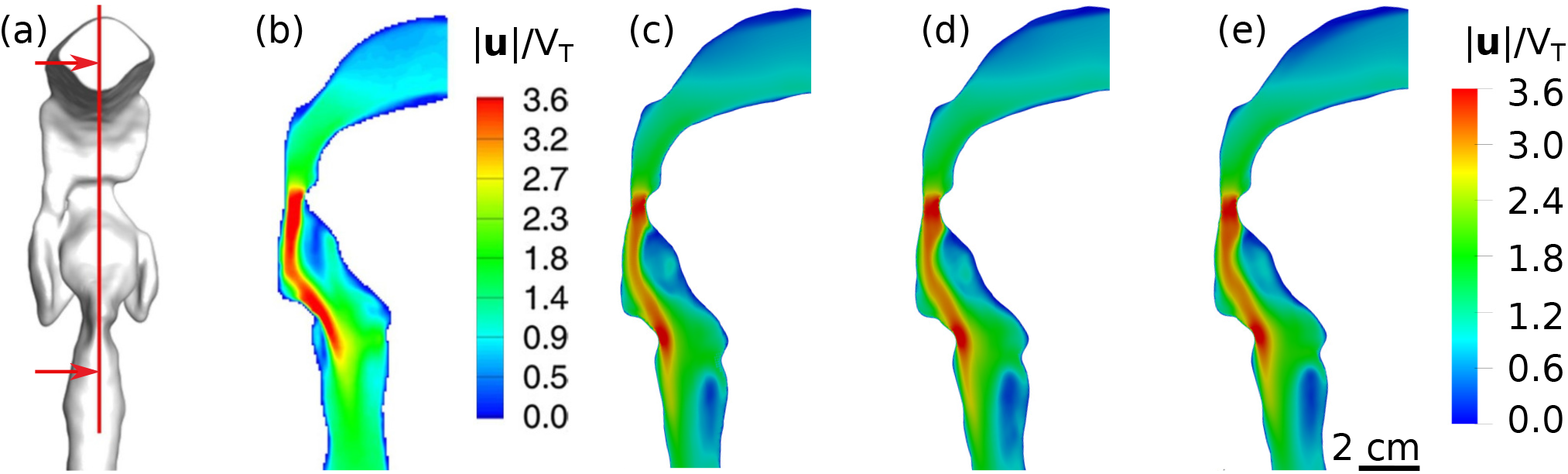
Comparison of single-phase flow in the upper airways to demonstrate mesh independence and independence of water and gas simulations at equal Reynolds numbers. (a) Location of shown contour (red) in domain (Banko et al., 2015, Figure 5), with arrows indicating direction of view for comparison of the normalised velocity magnitude contours of: (b) experimentally obtained water flow (Banko et al., 2015, Figure 5), and numerically obtained water flow on a (c) moderate grid density and (d) fine grid. (e) Numerically obtained airflow on a fine grid.

The resemblance in flow structure continues in the lower airways, from both coronal and axial views as given in Figure 4 (see Banko et al. (2015) for further comparison). The slight separation seen at the left bronchus (right side of (a)) agrees well with the experimental data. Contours in the axial region (shown in (b)) agree well in shape, accurately capturing recirculation and asymmetrical flow. However, velocity vectors shown in C-C’ and D-D’ differ from the published results. This likely stems from differences in time-averaging of such a sensitive parameter in highly unsteady flow. Vectors in B-B’ and E-E’ match the experimental data well. Qualitatively comparing numerical and experimental velocity fields (Banko et al., 2015) verifies that the single-phase flow configuration is suitable to capture the flow patterns present. To quantitatively validate the solver, we compare the relative bulk velocity (*U*/*V*_*T*_). Banko et al. (2015) gave this as 1.03 in the trachea, matching our solver exactly. In the left main bronchus, we had a value of 0.67, differing from the experimental results of 0.70 by 4.3%. In the right bronchus the numerical and experimental relative bulk velocity was *U*/*V*_*T*_ = 0.88 and 0.87, respectively (relative error 1.2%). These relative bulk velocity comparisons at key cross sections of the airway validate that our solver can reproduce the *in vitro* respiratory velocimetry measurements of Banko et al. (2015) to an error below 5%.

**Figure 4:**
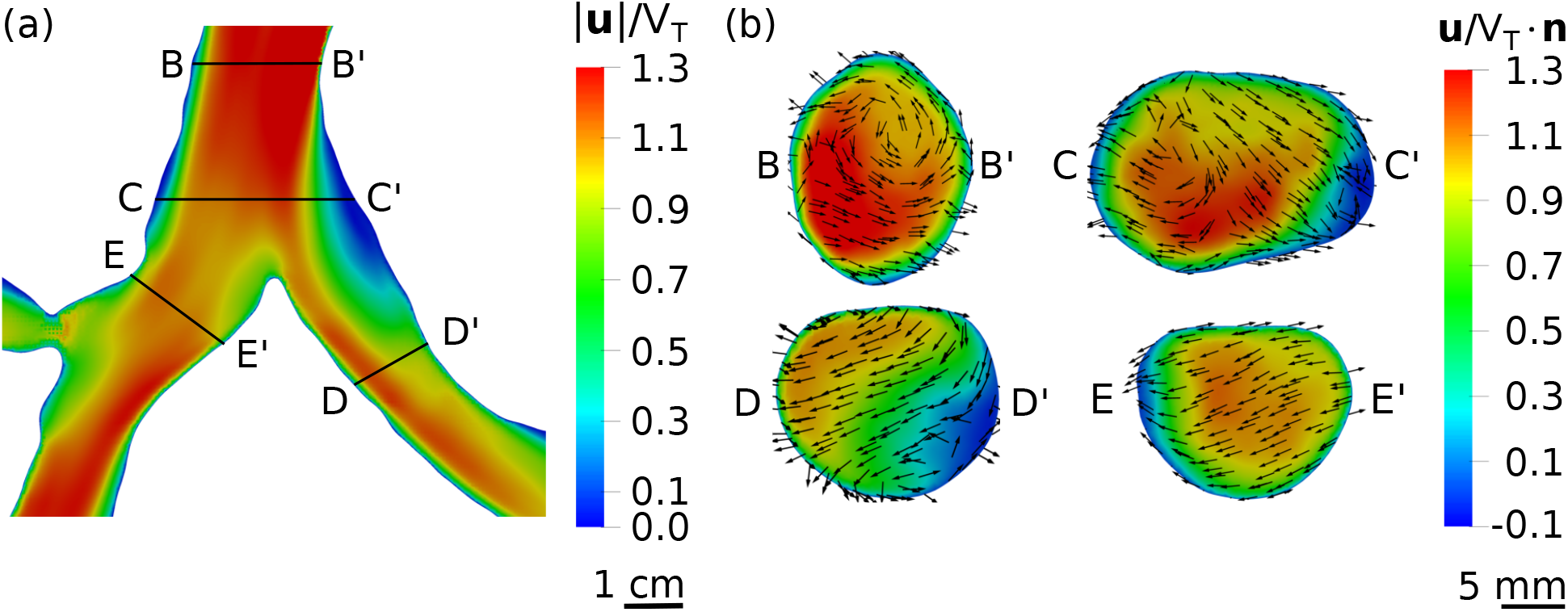
Time-averaged water velocity field at the first bifurcation, structured the same as presented in experimental study (Banko et al., 2015, Figure 9) for ease of comparison. (a) Normalised velocity magnitude at the first bifurcation from the coronal view, and (b) the velocity magnitude normal to the flow at various axial cuts shown in (a).

### 3.2. Effect of parameter variations

To model inhaler inhalation the simulation’s sensitivity to the dosage simulated, van der Waals forces and particle-wall lubrication forces were evaluated. The influence of each of these were examined by their influence on dosage deposition.

As physical parameters were varied, one can observe that deposition distinctions are mostly minor, as deposition changes across each lobe were all under 1.5% of the total dosage (Figure 5). When increasing the number of particles the only change came at *N*_*p*_ = 1, 000, 000 (Figure 5c), as the throat deposition rose by 2%, in all other regions the difference was less than 1%. For this reason 200, 000 particles was chosen for the remaining simulations to reduce simulation times. Although we have purposely underestimated *N*_*p*_, the number of particles should not be chosen arbitrarily. Instead the sensitivity of the model to this parameter should be included as an early set of simulations in all studies where the true payload cannot be simulated. When including particle cohesion and particle-wall lubrication (Figure 5h,i), deposition in the throat due to particle-wall lubrication forces rose by 13% of the total dosage. This effect is reduced by 4% when modelling van der Waals forces, likely explained by the agglomeration of particles providing additional inertia (increasing *St*_*coll*_), thus reducing the energy lost due to lubrication. This also explains why minimal changes are observed in Figure 5(d – f).

**Figure 5:**
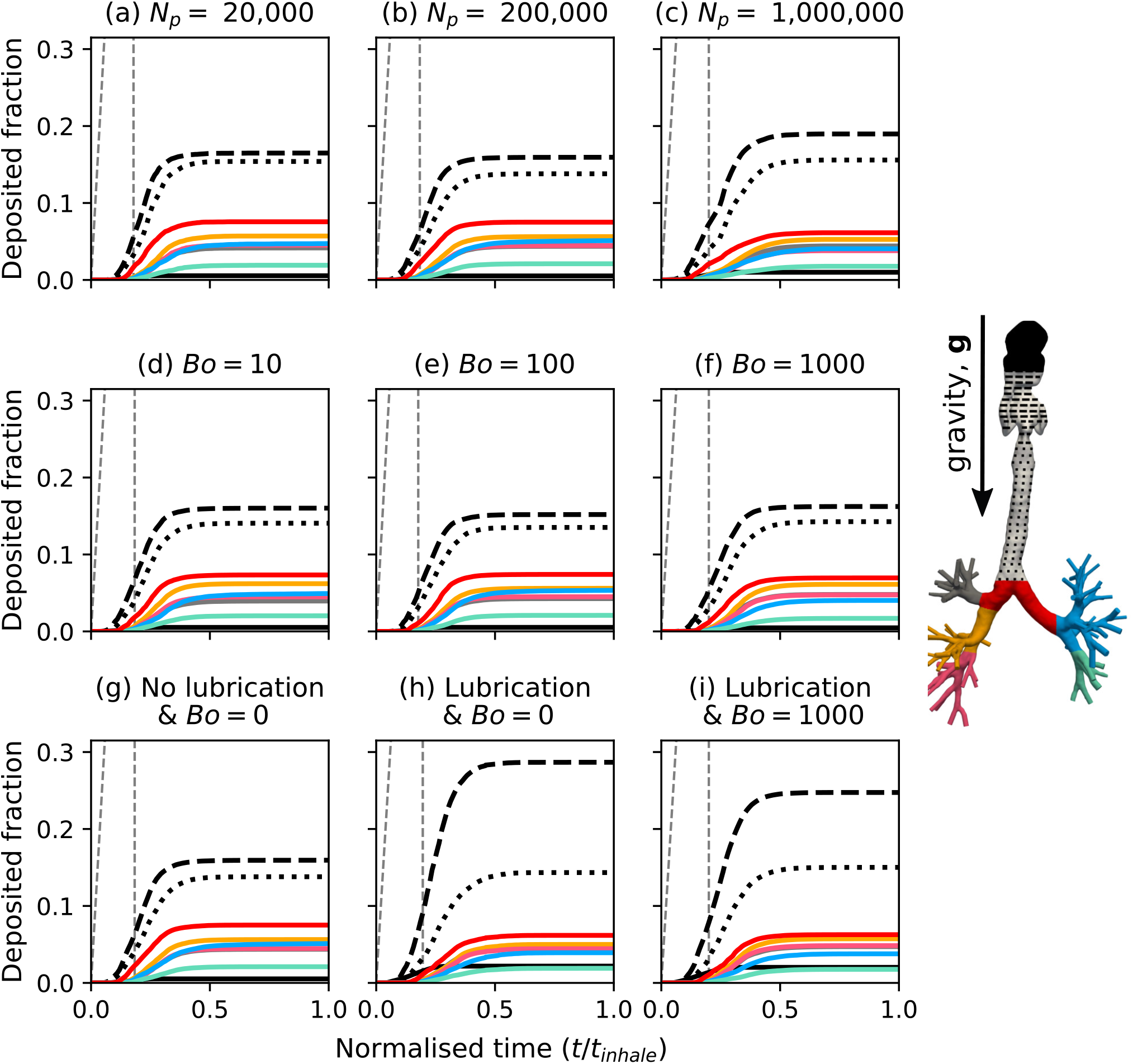
Dosage deposited within each region of the airways with time, for the parameter variations given in Table 2. (a, b, c) Compares deposition with simulation of various particle numbers. Remaining rows present results using *N*_*p*_ = 200, 000. (d, e, f) Shows deposition with variation of the van der Waals force in relation to the dimensionless quantity *Bo*. (g, h, i) Compares deposition with inclusion of lubrication forces and van der Waals particle-particle interactions. Each line is representative of an airway region, corresponding to the coloured airway included in the left-most plot of each row. An additional grey, dashed line is included to show the rate at which particles enter the system from the inhaler.

The parameters taken forward to evaluate deposition variance in the diseased patients were: a dosage of 0.126 µg (*N*_*p*_ = 200, 000 at *d*_*p*_ = 10 µm), a Bond number of 1000, and particle-wall lubrication forces. Time-varying breathing profiles were also applied (Figure 2). Although particle count is underestimated, the error is within 5%, for a computation time reduction of 62% (*N*_*p*_ = 200, 000 took three weeks, whereas *N*_*p*_ = 1, 000, 000 took eight). These parameters were considered to model the particle behaviour accurately, with a small sacrifice made to reduce computational cost.

### 3.3. Inter-patient variation

To evaluate the influence of airway shape on deposition we compare regional deposition for each patient in Figure 6. To understand the influence of breathing profile we also include the healthy patient with a healthy inhalation in Figure 6. Due to the dominance of upper airway deposition, it is difficult to visualise differences in central airway deposition. When viewing deposition as a logarithm the behaviour in the lobes can be analysed with greater ease (Figure 6). Across the five simulations the dosage was mainly deposited in the external airways (Figure 6), as deposition here was always greater than half of the payload. Given the size of the particles compared to the distribution used in inhalers, a large deposit here was expected. For the most constricted external airways (Figure 6c, d) the deposition fraction was 0.95 and 0.85, respectively. Deposition in the cystic fibrosis (dilated throat) patient’s trachea dropped by 13.4% relative to the total dose (Figure 6d, e). There is an external airway deposition rise of 1.9, 1.7 and 1.5 times when comparing the healthy patient with exacerbation breathing against the cancer and cystic fibrosis patients (constricted and dilated throats), respectively.

**Figure 6:**
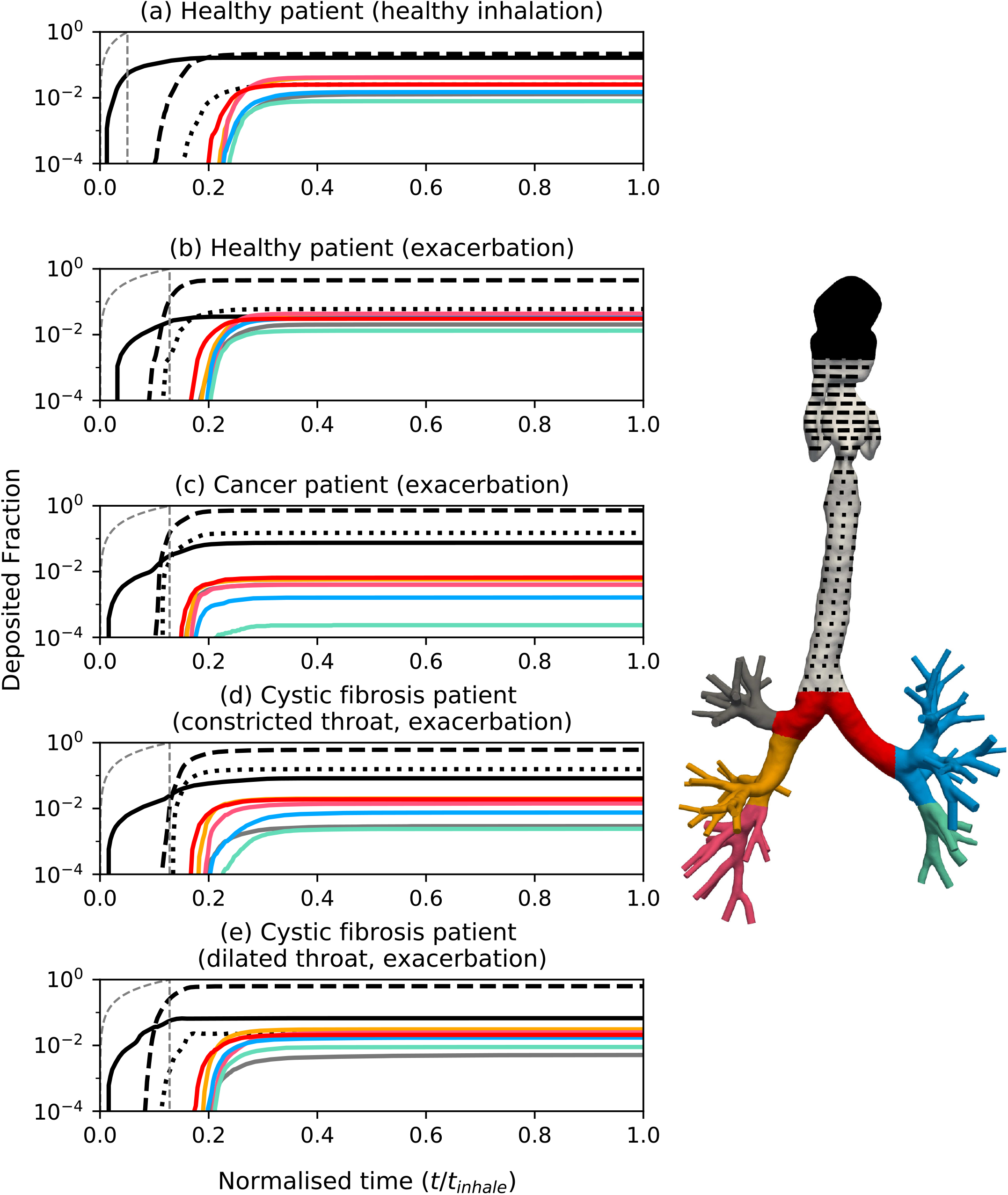
Comparison of regional deposited particles in three diseased model airways throughout a breathing cycle (Figure 2). Deposition is shown on a log scale to allow closer analysis of lobar deposits. Time has been normalised and patient illnesses described each plot. Line colours correspond to region in the coloured, generic airway tree on the left. Rate of particle injection to the domain given by the grey, dashed line.

We compared deposition in the external airways for the healthy patient, lung cancer patient and cystic fibrosis (with dilated throat) at *d*_*p*_ = 10 and 4 µm (Figure 7a, b). With a healthy breathing profile and 10 µm particles (Figure 7a), external airway drug deposition lowers by 23% of the dose for the healthy patient, relative to the deposition with rapid breathing. The external airway deposition is lowered by 43% relative to the deposition during an exacerbation for the cystic fibrosis patient (Figure 7a and Supplementary Video). Similarly, the deposition is lowered by 48% for the lung cancer patient (Figure 7a), relative to during an exacerbation. For the 4 µm particles (Figure 7b), the upper airway deposition did not change for the cystic fibrosis and lung cancer patients when comparing deposition under the two different inhalations. Upper airway deposition in the healthy patient lowered from 30% of the total dose to 22% when modelling the healthy inhalation.

**Figure 7:**
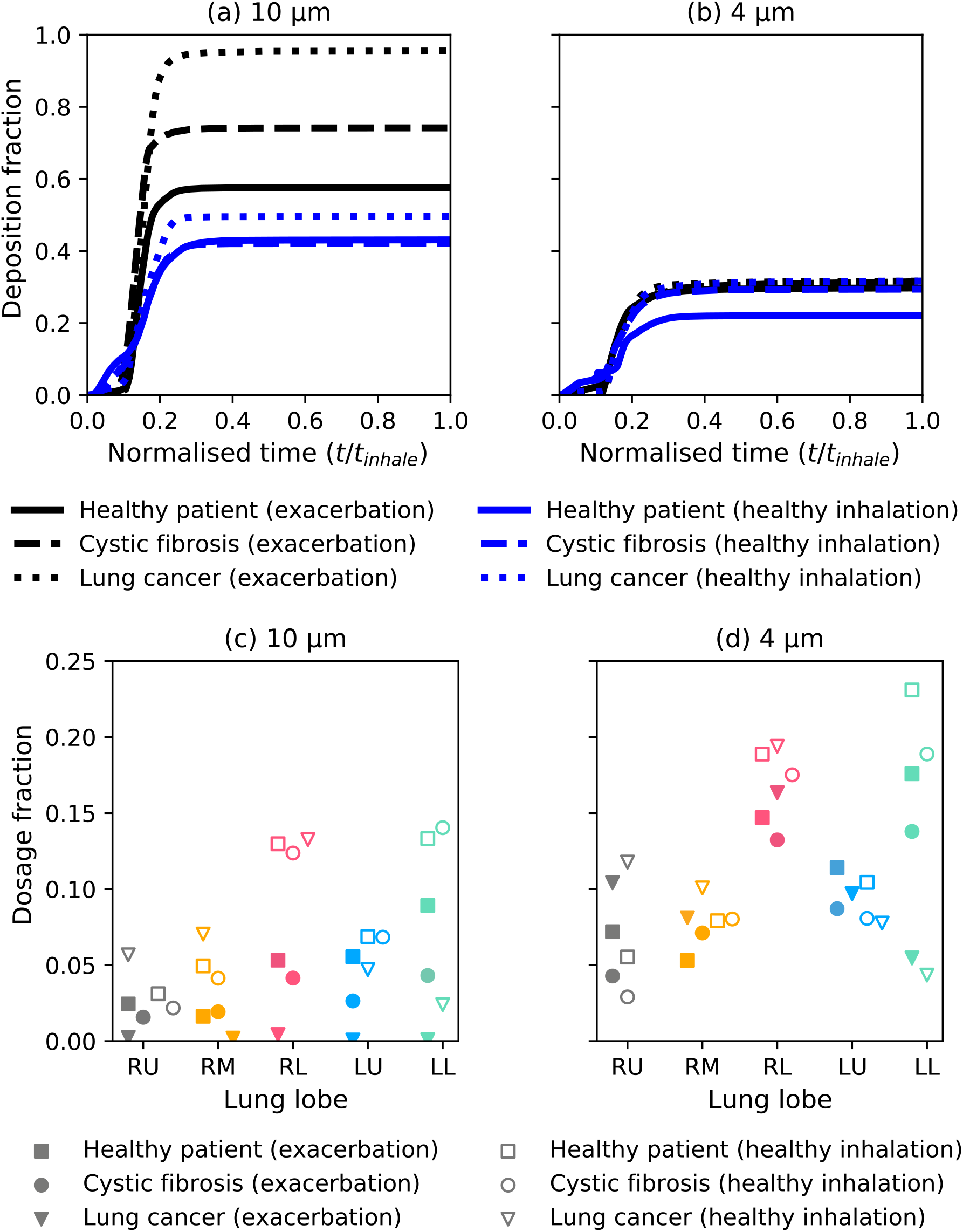
Comparison of breathing profiles for 10 µm, and 4 µm particles. The top row compares external airway deposition (mouth to first bifurcation). The bottom row compares lobar distribution of drug exiting the domain outlets to reach the deep lung. Line style is consistent with each patient. Black lines denote deposition with an exacerbation breathing profile and blue lines denote deposition with a healthy breathing profile. Solid markers correspond to drug reaching the deep lung with an exacerbation breathing profile and hollow markers correspond to a healthy breathing profile. Adapted from Williams et al. (2021) with permission of RDD Online LLC.

To compare the influence of airway shape and inhalation profile on the drug reaching the deep lung, we show the fraction of the total dose that exits the outlets of our CT-domain to the deep lung (Figure 7c, d). Reducing the particle size increased the dosage reaching the deep lung by 15 - 49% of the total dose across the three patients (Figure 7d). The largest increase (49% of the total dose) was in the lung cancer patient with the exacerbation breathing profile. In the 10 µm simulations, the dose reaching the deep lung increased by between 16 - 32% of the total payload (25% mean) for a healthy breathing profile compared to the exacerbation breathing profile (Figure 7c). The increase of 4 µm particles reaching the deep lung with a healthy breathing profile was 3.3 - 9.6% (7% mean) of the total dose (Figure 7d). As particles are known to distribute asymmetrically across the lungs (Lambert et al., 2011), we compared the ratio of particles in the right lung to left lung from Figure 7c and d. We also compared the ratio of drug in the upper and lower lobes. The right to left lung ratio was 1.04 and 0.96 in the healthy patient during a healthy inhalation for 10 and 4 µm particles, respectively. In the cystic fibrosis patient during a healthy inhalation, the right to left lung ratio was 0.89 and 1.05 for 10 and 4 µm particles. During an exacerbation, the right to left lung ratio was 1.09 for both sets of particles. The drug distribution was most asymmetric for the lung cancer patient where the right to left lung ratio ranged from 2.29 to 5.62. The ratio particles in the lower lobes to upper lobes (neglecting the right middle lobe) ranged from 1.08 in the lung cancer patient (exacerbation, *d*_*p*_ = 4 µm) to 3.31 in the cystic fibrosis patient (healthy inhalation, *d*_*p*_ = 4 µm) with a mean value of 2.04. On average the lower to upper ratio was 38% larger with the healthy breathing profile than the exacerbation for *d*_*p*_ = 4 µm and 22% larger for *d*_*p*_ = 10 µm (mean for all *d*_*p*_ is 35%). The asymmetry of drug distributed in the lobes can be seen for the 10 µm particles in the cystic fibrosis patient with both inhalations in the Supplementary Video.

### 3.4. Dosimetry assessment

To evaluate local changes in deposition due to airway shape and inhalation profile, we compare the DEF for 10 µm particles (Figure 8). All simulations featured hotspots in mouth and throat which ranged from a maximum value of 116 for the lung cancer patient with a healthy inhalation to 430 for the cystic fibrosis patient during an exacerbation. The maximum DEF in the bronchial airways of the healthy patient and cystic fibrosis patient was 20 during a modelled exacerbation and 40 during a healthy inhalation (increased by a factor of 2). Similarly, the maximum DEF in the lung cancer patient’s bronchial airways increased from 3 during a modelled exacerbation to 45 during a healthy inhalation (increased by a factor of 15).

**Figure 8:**
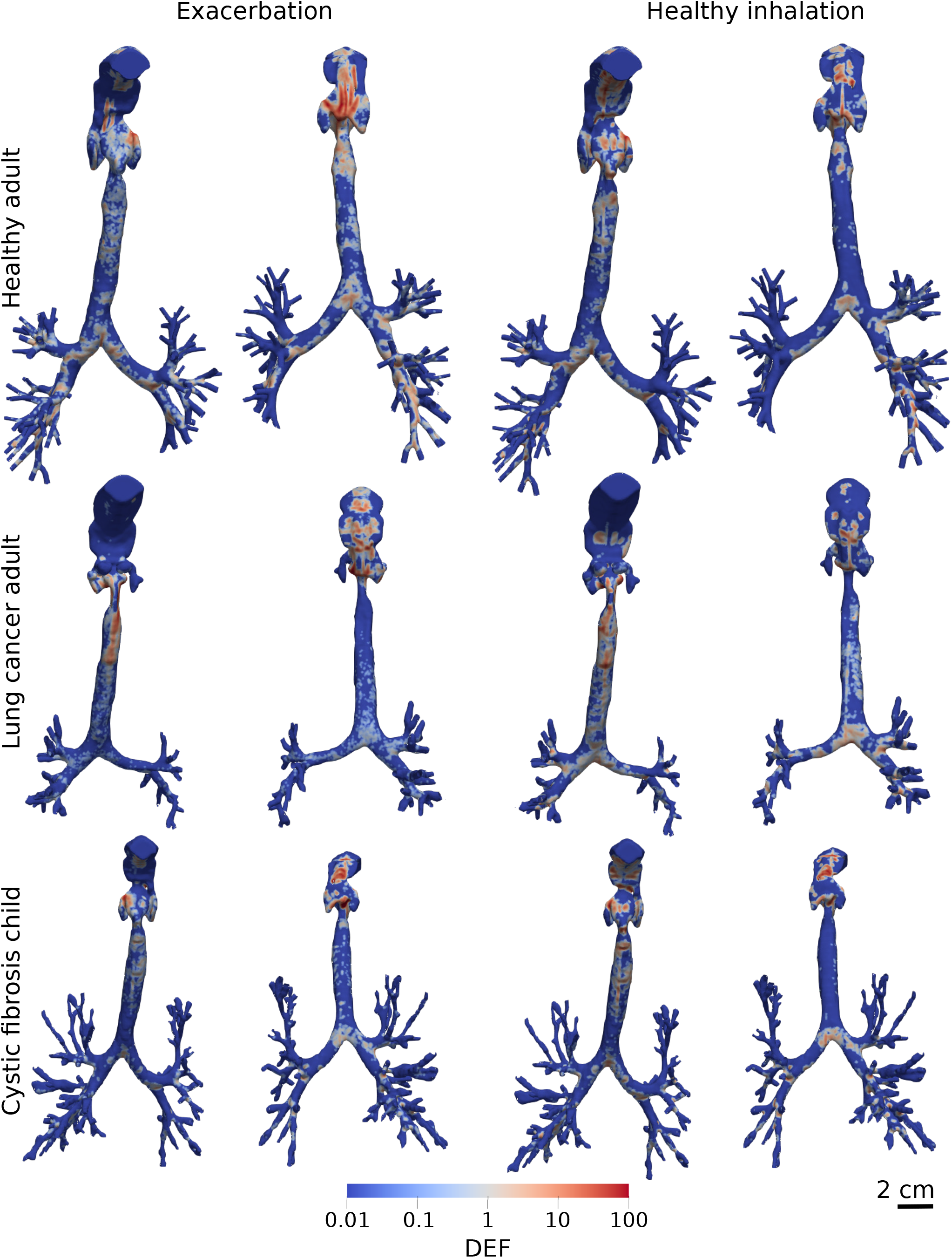
Concentration of 10 µm drug particle deposited on the airway wall of each model, normalised by total global deposition (DEF, Equation (6)). Hotspots visualised from the front (first and third columns) and rear (second and fourth columns) of each model. The maximum value (DEF = 100), describes an area with a drug concentration 100 times greater than the mean DEF of 1. Dosage is most concentrated in the throat as shown in Figure 6.

During an exacerbation the 4 µm particles are more uniformly dispersed than during a healthy inhalation (Figure 9). The maximum DEF in the bronchial airways increased from 44 during an exacerbation to 68 during a healthy inhalation in the cystic fibrosis patient (56% increase). Similarly, the maxmimum DEF in the lung cancer patient’s bronchial airways increased from 33 during an exacerbation to 46 during a healthy inhalation (43% increase). The maximum DEF increased from 32 during an exacerbation to 91 during a healthy inhalation in the healthy patient’s bronchial airways (190% increase). The airway surface area with a DEF value of at least one (DEF >= 1) increased by 89% in the cystic fibrosis patient in the 4 µm simulations compared to the 10 µm simulations. This surface area increased by 113% for the lung cancer patient compared to the 10 µm simulations.

**Figure 9:**
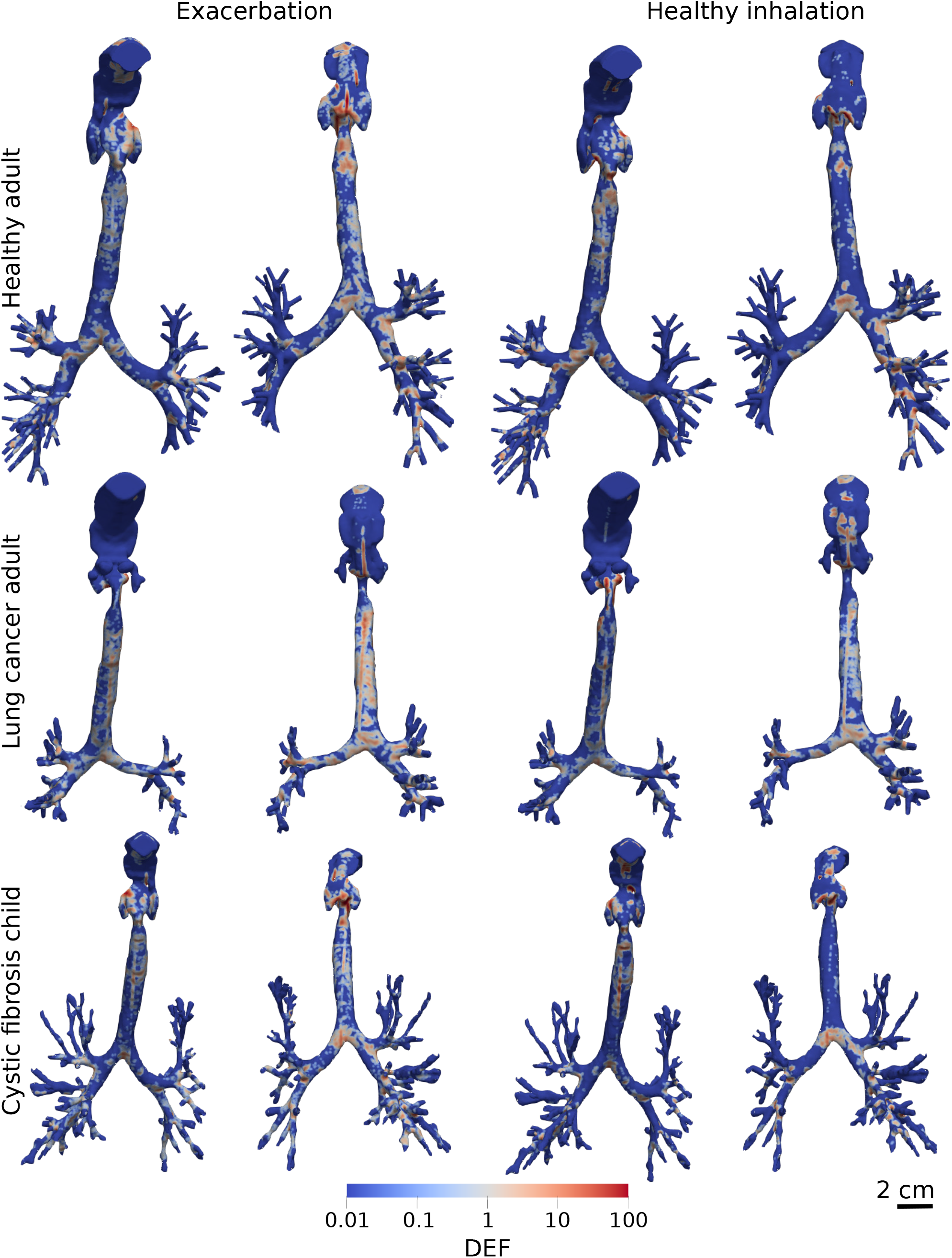
Concentration of 4 µm drug particle deposited on the airway wall of each model, normalised by total global deposition (DEF, Equation (6)). Hotspots visualised from the front (first and third columns) and rear (second and fourth columns) of each model. The maximum value (DEF = 100), describes an area with a drug concentration 100 times greater than the mean DEF of 1. Dosage is most concentrated in the throat as shown in Figure 6. Adapted from Williams et al. (2021) with permission of RDD Online LLC.

## 4. Discussion

In this study we aimed to evaluate deposition in patient-specific models during the rapid breathing of an exacerbation, compared to during healthy breathing. This allowed the investigation to consider ‘reliever’ inhalers. By investigating this in a small, diverse patient group we found patient-specific domains to be a necessity in future studies. For large particles (10 µm), there was a significant change in upper airway deposition with airway shape and breathing profile. Small particles (4 µm) showed less sensitivity to total upper airway deposition for all airway shapes and breathing profiles. This shows that small particles are optimal for improving drug penetration to the deep lung during exacerbation. Local deposition concentration (DEF) varied for all particle sizes, airway shapes and breathing profiles, even in cases with identical regional deposition. Therefore, we recommend that patientspecific models should be used to determine local concentration as this is a main driver of clinical outcome.

We used highly-resolved particle tracking (DEM) to allow us to include cohesive and collision forces. This was as cohesive van der Waals forces have been known to influence the transport of micro-scale particles (Gu et al., 2016*a*). Soft-sphere (treating particles as deformable) deposition studies have been implemented before but only to observe dynamics in simplified airways (Chen et al., 2012; Wang et al., 2017). We conclude that the computational cost of DEM is too large to be feasible for further study in broad patient populations with varying inhalation parameters and drug properties (particularly smaller particles). The results in Figure 5 suggest that cohesive and collision forces are of insignificance and may be replaced by some stochastic collision tracking model such as that of van Wachem et al. (2020). However, the computational cost of DEM also limited the dosage that could be modelled to 0.1 µg, which is 100 − 1000× less than a realistic dosage (≈ 100 µg). In reality, the effects of cohesive and collision forces may be more influential as the particles will be more densely concentrated. Also any effect of collision and cohesion will increase in juvenile patients due to their small airways creating a denser particle packing during inhalation. As suggested by Islam et al. (2020) the effect of particle-particle interactions should be investigated across many cases to understand their influence on respiratory drug delivery, but our study has provided a first step towards this. To use modelling to inform patient treatments an understanding of different particle forces and their change with drug properties is important.

Dosimetry analysis across the patients show noticeable differences in deposition in the trachea (Figures 8 and 9), attributable to transitioning from the fast-flowing, constricted throat to the expanding and curved trachea, of the cystic fibrosis and cancer patients. This agrees with single-phase simulations of Wei et al. (2017) that found flow to vary with orientation, as well as Bates et al. (2016) who showed the effect of pathological trachea curvature on gas pressure and energy loss. Also the change in relative orientation of the trachea to gravity present an additional factor to deposition changes. The healthy inhalation creates deposition hotspots primarily at the bifurcation points (Figures 8 and 9). This occurs as the effect of particle drift towards the wall of the trachea (turbophoresis) is naturally weaker in a lower Reynolds number flow (Bernardini, 2014), which makes particle deposits mainly at bifurcation points due to inertial impact (Zhang et al., 2018). However, the deposition of 4 µm was generally more uniformly dispersed on the airway wall during an exacerbation, than during a healthy inhalation (Figure 9). This occurs as the small particles have low inertia and are more influenced by turbulent secondary flows. As drug concentration across the airway surface is important in its dissolution into the tissue (Solomon et al., 2012), this provides a deeper understanding of the drug’s absorption than a simple deposition analysis. These results could be input to a model of the particle’s interaction with the tissue at smaller scales, as has been done by Olsson and Bäckman (2018) using 1D deposition data. Using CPFD for this could further inform clinicians in comparing potential treatment and delivery techniques for a patient.

When observing intra-patient drug delivery, the results presented show an uneven distribution across the lobes. There is a clear favour in transport to the lower lobes of the lung (Figure 7c, d). This is similar to observations made by Lambert et al. (2011), who used a single constant inlet velocity. However, our results extend this by showing that the number of particles in the lower lobes increases during a normal inhalation compared to an exacerbation. Patient-specific models could be used to attain a balanced dosage distribution, or target a specific region. This could allow for improved symptom relief through inhaler design informed by knowledge of regions sensitive to local inflammation (Barbu et al., 2011), allowing for more efficient devices. Additionally, cancerous regions of the lung could be targetted for chemotherapy using nebulisers (Tatsumura et al., 1993; Kleinstreuer and Zhang, 2003; Kleinstreuer et al., 2007). Clinicians could therefore predict and tailor the chemotheraputic agent delivery through patient-specific CPFD models to minimise radiation reaching undesired areas of the lung. As a course of chemotherapeutic treatment is given over a period of around three weeks (Wittgen et al., 2007), this fits well with a high accuracy, time-consuming modelling method such as CFD-DEM.

Limitations of this study included the absence of imaging data available for the throat and mouth of the child, meaning this region was taken from the other patients and scaled. This of course limits the specificity of the main deposition site. However by simulating with the two available throat geometries merged to the child’s trachea we can see that the deposition in the mouth and throat is unaffected (only changing 1%). The only differences are observed downstream in trachea deposition (Figure 6). Therefore, to neglect this region completely would harm the accuracy of the results downstream as flow generated in the upper airways heavily impacts downstream behaviour of both the gas and the particles (Figure 4). However, adding this region from adult patient to a child does not take into account the maturational effects of adolescence on the airway, which may impact deposition here. In future studies, images containing the mouth and throat are needed to ensure this important section is patient-specific.

An additional limitation is that patients lying down in scans may have slightly different airway shape and orientation when standing or sitting upright (Jan et al., 1994), as is a typical posture when taking inhalers. If using image-based CPFD to recommend treatments, the patient images should be consistent with their typical posture when using their treatment where possible.

Only modelling the small number of airway bifurcation levels visible in the CT scan limits the model to only providing deposition information about the upper and central airways. To understand drug delivery in the smaller more distal airways the particles ‘exiting’ from our model’s outlets could be coupled to analytical 1D models (Kuprat et al., 2020; Koullapis et al., 2019). These models predict deposition based on particle size, estimated airway length and diameter, and flow rate. This would allow for a coarse prediction of drug delivery and efficacy in the targeted small airways.

Assumptions included the simulation of a uniform particle distribution. A uniform distribution is commonly used to research aerosol physics experimentally (Usmani et al., 2005, 2003), although does not directly represent the varying size distribution of inhaler particles (Dolovich, 1991). This is due to the small range of particle sizes (geometric standard deviation below two (Mitchell et al., 2003)) in a real device, meaning *St* < 1 in all cases, therefore the particles will exhibit high sensitivity to flow changes in both polydisperse and monodisperse flows. Deposition results should not be appreciably different as no transport characteristics are changed by this approximation. This allowed patient morphology differences to be evaluated in a simpler manner, without sacrificing accuracy.

Our simulations did not account for the effect of turbulent dispersion on particle transport. We used the interpolated fluid velocity to calculate the drag force acting on a particle, which neglects the influence of subgrid fluctuations in the motion of the gas phase. The low inertia of aerosol particles (*St* << 1) means that they are sensitive to small changes in the gas velocity. Filtering out these subgrid fluctuations has been shown to alter inertial particle (*St* >= 1) segregation towards the wall and the resultant deposition in simple geometries such as channel flow (Marchioli et al., 2008). Using a stochastic model for the instantaneous velocity of fluid ‘seen’ by the particle has been shown to better predict inertial particle concentration (Innocenti et al., 2016), but has not been investigated in LES-based airway deposition modelling. Additionally, such a model is currently not available in OpenFOAM. Our future study will investigate the effect of turbulent dispersion on drug deposition in patient-specific airways.

We softened the particles to increase the DEM timestep and lower the computational cost. Particle softening is standard in DEM simulations to permit a larger DEM timestep as the particle’s collision happens over a longer period due to a lower stiffness. We mitigate an impact of increased van der Waals forces by including the cohesion model of Gu et al. (2016*a*). This model makes particle cohesion independent of softened particle stiffness, and therefore increases our simulation timestep from *t*_DEM_ = 0.8 ns to 25 ns for *d*_*p*_ = 10 µm. This reduction of real particle stiffness (*k*_*R*_) to a softer stiffness (*k*_*S*_) has been shown to have negligible effect on fluid hydrodynamics and particle cohesion (Gu et al., 2016*a*; Ozel et al., 2017). For dry-powder inhalers, where the particle transport is primarily dependent on cohesion, Gu et al. (2016*a*)’s model may not be applicable. However, this is not the case for the mildly cohesive particles modelled in this study (Gu et al., 2016*a*). The use of this model therefore makes the effect of softening particles negligible in our simulations. This allows us to use DEM to model inhaler particle-particle and particle-wall interactions with feasible computation times.

## 5. Conclusion

We have performed patient-specific inhaler deposition simulations across three diverse patients during exacerbating inhalation conditions compared to a healthy breathing profile. Our results showed that during an exacerbation less of the drug reaches the deep lung for 10 and 4 µm particles. For 10 µm particles, the total upper airway varied strongly with inhalation, but the change was small for 4 µm particles. The ratio of drug in the lower lobes to upper lobes of the lung was 35% larger during healthy inhalation than an exacerbation, showing enhanced asymmetry during slower inhalation. For both particle sizes the size, location and intensity of local deposition hotspots varied with patient inhalation and airway shape. This demonstrates that image-based models are needed to predict therapeutic outcome in respiratory CPFD studies. Image-based models should be combined with inhalation profiles that represent various symptoms such as an exacerbation to optimise patient treatments.

## Data Availability

Simulation data available upon request.

## Acknowledgements

For their early involvement and insight into the issue faced by patients, the authors would like to thank Elisabeth Ehrlich and Olivia Fulton. Also Carol Porteous (Patient Public Involvement Advisor) who arranged our contact.

The authors thank Prof. Vicki Stone from Heriot-Watt University for fruitful discussions. The authors also thank Dr Filippo Coletti from University of Minnesota for providing the surface file of the healthy patient’s segmented airways that enabled our model validation and was used to compare deposition across the patients. AO thanks Prof. Sankaran Sundaresan from Princeton University for his support and fruitful discussions. JW thanks Dr Rudolf Hellmuth from Vascular Flow Technologies for fruitful discussions.

JW was funded by an Institution of Mechanical Engineers Postgraduate Masters Scholarship 2018, a Scottish Funding Council Masters fee scholarship 2018, and a Carnegie-Trust for the Universities of Scotland PhD scholarship 2019.

## 6. Conflict of Interest

The authors declare that they have no known competing financial interests or personal relationships that could have appeared to influence the work reported in this paper.

## Appendix A. Gas and solid-phase modelling

Simulations in this study solve fluid transport through the respiratory system on a Eulerian grid with Lagrangian particle tracking using the discrete element method (DEM). We solve the volume-filtered mass (A.1) and momentum (A.2) balance equations at each finite-volume cell. Here they are presented in terms of the volume-filtered variables, accounting for particle interactions as derived by Anderson and Jackson (1967) and Capecelatro and Desjardins (2013), with overbars denoting filtered terms and bold, lower- and upper-case characters describing vectors and second order tensors, respectively,

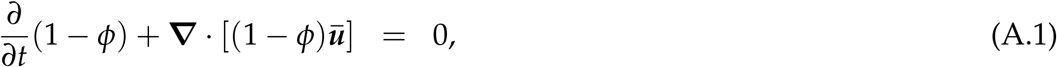

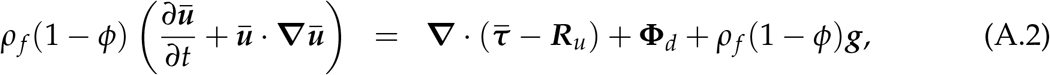

there *ū* is the filtered local velocity vector, 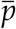 is the filtered local fluid-pressure, *ρ* _*f*_ is the fluid density, *ϕ* is the particle volume fraction, ***g*** is gravitational acceleration (direction shown in Figure 5), the force caused by interaction with the discrete phase is −**Φ**_*d*_, ***R***_*u*_ is the sub-grid stress from filtering, modelled using a dynamic Smagorinsky model (described in Section 2.2). 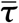 is the filtered fluid stress tensor, composed of the fluid pressure gradient 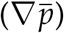, the deviatoric viscous stress tensor (labelled below), and an additional term arising from filtering of sub-grid velocity fluctuations (***R***_*µ*_) (Capecelatro and Desjardins, 2013)

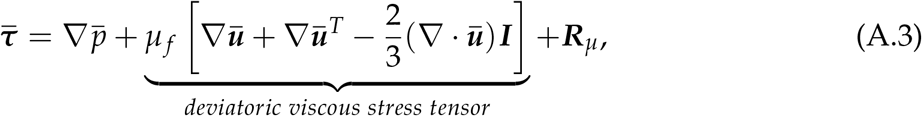

where ***I*** is an identity tensor and ***R***_*µ*_ is the term arising from filtering velocity gradients. In this study we dismissed ***R***_*µ*_ to be included in a later study, and the deviatoric part of the stress tensor due to its minor influence on gas-solid flows in comparison to ***R***_*u*_ (Agrawal et al., 2001). **Φ**_*d*_ is dependent on the interaction force between particles and fluid (***f*** _*f* →*p,i*_) of all particles within a cell volume (V_cell_) by, 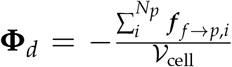 (Anderson and Jackson, 1967; Capecelatro and Desjardins, 2013; Ozel et al., 2017). For a particle *i*, this is related to the filtered fluid stresses and the particle’s drag by 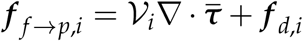 where ***f*** _*d,i*_ is the drag force, taken from Beetstra et al. (2007).

Newton’s equations of motion are used to track particle linear motion (A.4) and angular motion (A.5) (Cundall and Strack, 1979; Capecelatro and Desjardins, 2013), by

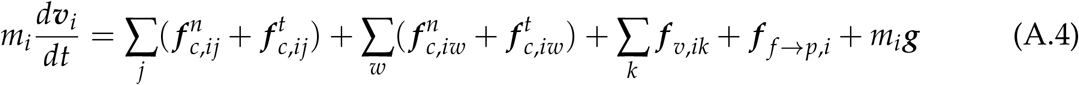

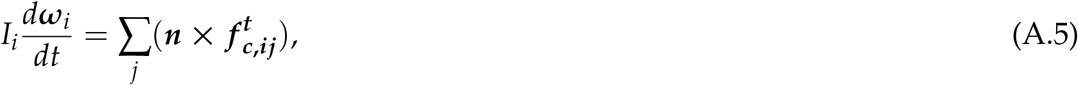

where *m*_*i*_ is the mass of a particle, *i, v* and *ω* are the particle’s translational and angular velocity, respectively, ***f***_***c***_ is the contact force from a particle-particle collision (subscript _*ij*_), and particle-wall collision (subscript _*iw*_), in the normal and tangential directions shown by sub or superscript *n* and *t*, respectively. van der Waals forces are shown with (.)_*v*_. Angular momentum (A.5) from inter-particle collisions depends on outward unit normal vector from particle centre to the point of collision, ***n***, and the tangential contact force 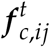. Particle contact forces are solved here using a linear spring-dashpot model (Cundall and Strack, 1979; Capecelatro and Desjardins, 2013),

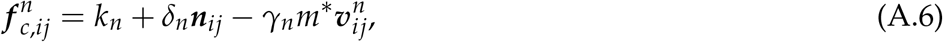

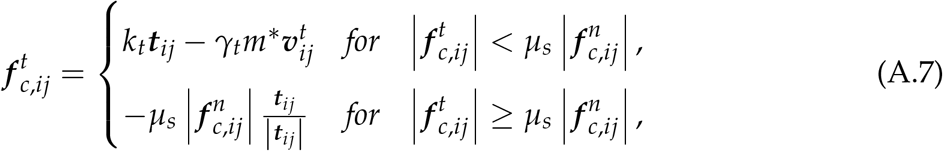

where *k*, is the particle’s spring constant with *k*_*t*_ = 2*k*_*n*_/7 (Matuttis et al., 2000), *δ* is the particle overlap distance, *γ* is the viscous damping coefficient. *γ*_*n*_ is calculated from the coefficient of restitution 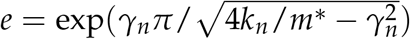 (Gu et al., 2016*b*). *γ*_*t*_ = 2*γ*_*n*_/7. Effective particle mass is *m*\* = *m*_*i*_*m*_*j*_/(*m*_*i*_ + *m*_*j*_), in particle-wall collisions *m*\* = *m*_*i*_, as one radius is assumed infinite (Gu et al., 2016*a*); *µ*_*s*_ is the sliding coefficient; ***t***_*ij*_ represents the tangential displacement due to a collision, found from the integral of its velocity component.

## Notes

### Competing Interest Statement

The authors have declared no competing interest.

### Funding Statement

JW was funded by a Carnegie-Trust for the Universities of Scotland PhD scholarship (2019-2022), an IMechE Postgraduate Masters scholarship (2018) and a Scottish Funding Council Masters fee scholarship (2018).

### Author Declarations

This research was approved by Heriot-Watt University Engineering and Physical Sciences Ethics Committee (ID: 2020-0500-1452).

### Summary of Updates

Updated results in Sections 3.3 and 3.4.

